# Psychiatric symptoms and syndromes transcending diagnostic boundaries in Indian multiplex families: The cohort of ADBS study

**DOI:** 10.1101/2020.01.06.20016543

**Authors:** Vanteemar S Sreeraj, Bharath Holla, Dhruva Ithal, Ravi Kumar Nadella, Jayant Mahadevan, Srinivas Balachander, Furkhan Ali, Sweta Sheth, Janardhanan C. Narayanaswamy, Ganesan Venkatasubramanian, John P. John, Mathew Varghese, Vivek Benegal, Sanjeev Jain, YC Janardhan Reddy, ADBS Consortium, Biju Viswanath

## Abstract

Accelerator program for discovery in brain disorders using stem cells (ADBS) is an ongoing longitudinal study investigating the neurobiological aspects of five psychiatric disorders (Alzheimer’s dementia, bipolar disorder, obsessive-compulsive disorder, substance use disorder or schizophrenia) in India. The study uses several techniques (brain-imaging, psychophysics, neuropsychology, next-generation sequencing, cellular models), and in-depth clinical assessments in a longitudinal cohort from multiple-affected families. This article explores the frequency of manifestations of different psychiatric symptoms and syndromes in the participants and their relatives from the first wave of this study (August 2016 to October 2019). We screened 3,583 families and enrolled 481 families (1406 participants; 773 affected with any of the 5 disorders, and 633 relatives). The participants had a high familial prevalence with nearly a third of FDRs affected. Though similar disorders aggregated, the majority (61%) of the families had dissimilar diagnoses among members. Moreover, 15% of affected participants had two or more co-occurring syndromes. Diverse cross-cutting symptoms, unrestricted to the index syndrome, were observed in participants across diagnostic categories. The pattern and extent of co-occurrence validate the need for a transdiagnostic approach. The repository of biomaterials as well as digital datasets will serve as a valuable resource for the larger scientific community.

## Introduction

Psychiatric disorders are a significant cause of morbidity and ill health. A better understanding of the basic mechanisms of pathobiology that contribute to these syndromes is essential for improved interventions^1^. Recent findings, using diverse strategies ranging from epidemiology, and descriptive psychopathology, to pathophysiology and genetics, suggest that the diagnostic categories (of the various syndromes) may not be distinct and that transdiagnostic studies are thus necessary for brain-based disorders^2-17^. The transition in research from traditional categorical approaches to brain-based dimensional approaches has been proposed by the Research Domain Criteria (RDoC) project following suggestions by the National Institute of Mental Health^18, 19^. This approach is further validated by large-scale, cross-disorder genetic studies, that show a significant genetic overlap between multiple psychiatric disorders^3, 20-23^. Such overlaps have also been found in terms of brain markers in recent transdiagnostic studies^8, 24^. The presence of overlaps does not necessarily mean an absence of specific factors, though attempts to identify these have had equivocal success^17^.

One of the reasons for difficulty in finding disorder-specific factors is that the single-disorder, as well as cross-disorder, studies have generally not paid adequate attention to the familial risk of other disorders. The risk of psychiatric conditions is known to be transmitted in families, perhaps through a combination of both genetic and environmental factors. Individuals from families with multiple affected members could have a dense accumulation of the biological and environmental factors^25^ that contribute to risk. The amount of overlap of symptoms and syndromes in the members of the multiplex families thus needs to be better documented and investigated.

Most of the large-scale cross-disorder efforts in this direction are situated in high-income countries in Europe and North America^26^. The Accelerator program for Discovery in Brain disorders using Stem cells (ADBS) is an effort to bridge this gap using a transdiagnostic longitudinal cohort of multiple-affected families in India^27^. Large family sizes and high levels of endogamy in India provides a unique and valuable source for family-based studies in understanding the pathogenesis of complex mental disorders^28-31^. We aimed at evaluating the frequency of occurrence of different symptoms and syndromes in relatives of those with severe mental illness. The major psychiatric disorders do not necessarily ‘breed true’, and it is often observed that index probands have a family history of different psychiatric illnesses^27, 32-35^. The current analysis thus also aimed at evaluating the aggregation patterns of different disorders in multiplex families.

## 1. Methods

This analysis draws on data from the ADBS project, an ongoing longitudinal collaborative by the National Institute of Mental Health and Neurosciences (NIMHANS), the National Centre for Biological Sciences (NCBS) and the Institute for Stem Cell Science and Regenerative Medicine (InStem), which began in August 2016 ^27^. Participants were identified from the health services of NIMHANS. Individuals with a diagnosis of substance use disorder (SUD), bipolar disorder (BD), Alzheimer’s dementia (AD), obsessive-compulsive disorder (OCD) or schizophrenia (SCZ) under clinical care at the hospital were screened for an additional presence of a first-degree relatives (FDR) with either the same or any of the other four disorders. A total of 3,583 patients (from August 2016 to October 2019) attending the adult psychiatry services and different specialty psychiatry clinics (Geriatric clinic/center for addiction medicine/Schizophrenia clinic/Obsessive-compulsive disorder clinic) of the NIMHANS were screened. The study population was selected from a combination of modal instance sampling (multiplex family), diversity sampling (representation of 5 different disorders) and convenience sampling (from a tertiary health care center) methods. Identified families with multiple affected individuals were invited to participate with as many members (affected as well as unaffected) from each family as possible. Written informed consent was obtained from each participant and the proposal was approved by the institutional ethics committee. A detailed pedigree was drawn and the family interview for genetic studies (FIGS) based screening for psychiatric disorders was carried out. This was performed by an independent interview of at least two informants by a trained mental health professional (psychiatrist/psychiatric social worker/clinical psychologist). All psychiatric diagnoses were corroborated by two trained psychiatrists with ascertainment using the Mini International Neuropsychiatric Interview (MINI)^36^ and in accordance with DSM-IV-TR ^37^.

### 1.1. Baseline characteristics of the participants

The biographic and clinical profile of the initial 1406 participants from 481 multiplex families is described in this paper. Diagnoses within the main DSM-IV category of SCZ and related disorders were subsumed under SCZ and both bipolar I & II disorders were considered as BD. Individuals who had met criteria more than one of the psychiatric diagnoses were grouped as “Complex severe mental illness (Complex SMI)”. Schizoaffective disorder, bipolar type, was considered as Complex SMI and analyzed as individuals having phenotypes of both SCZ and BD. Dependence on any of the psychotropic substances in their lifetime, excluding nicotine, were considered as SUD. Some of the relatives who were found to have psychiatric disorders other than the five disorders under study were grouped as “other psychiatric disorders” and those without any axis-I psychiatric condition were regarded as “unaffected relatives”.

DSM5 self-rated Level-1 cross-cutting symptom measure-adult^38^ was used to elicit dimensional psychiatric symptom profile of the participants (n=1373; missing=33 which included 25 participants aged less than 18 years) in the past 2 weeks. It measures the frequency of the symptoms in the past two weeks and those symptoms crossing the threshold were counted as significant symptoms. The proportion of individuals with each significant symptom prompting clinical attention was calculated across each diagnostic category.

### 1.2. Clinical profile of the families

The identified families could be further defined as “families with similar disorders” (all affected individuals have the same syndrome) or “families with dissimilar disorders” (members may have any of the five disorders in the family) based on the FIGS assessment of FDR and second-degree relatives (SDR) of each participant in the study. The prevalence of psychiatric morbidities in FDRs and SDRs of the study participants from multiplex families was calculated with FIGS based on family history data.

As multiple members from a single-family are included as probands, the same relatives would be included independently multiple times in the series. For example., if two siblings participated in the study, their parents would be considered twice for the prevalence estimation. To overcome the unequal probabilities in the sample design, a complex sampling design statistic in IBM® SPSS® Statistic version 23.0 (IBM® corp,) was used for the analysis of prevalence rates. Each family was considered a cluster and the estimates were weighted for the number of participants from each family. A simple random sample design was assumed within the clusters for the standard error calculation with finite population correction^39, 40^.

Lifetime prevalence in relatives was calculated as the proportion of relatives who have ever had a psychiatric disorder at any time in their life up to the time of assessment. Similar to Weinberg’s shorter method, relatives who were below the age at risk for developing psychiatric conditions (<13 years) were excluded from this calculation^41^. As individuals of all the age groups are potentially at risk for developing AD in their lifetime, we did not use the age correction for at-risk age and above-risk age. Due to the unavailability of data, niece/nephew and grandchildren were not considered in the SDRs.

The prevalence of each of the five disorders in the relatives was calculated separately^42^. Also, the transdiagnostic prevalence was calculated for the presence of any of the five diagnoses within the family. They were described across participant’s diagnoses. Thereafter, the prevalence rates (P) and prevalence ratio (PR) of having similar and dissimilar diagnoses were calculated for each of the five diagnoses. An example formula for BD is shown below:

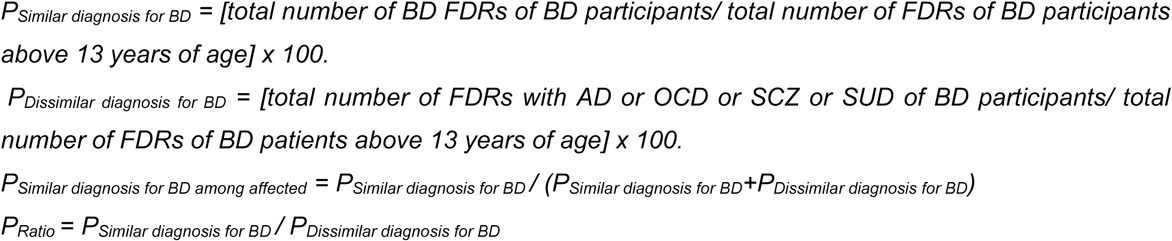

Similarly, P and PR were calculated for the psychiatric conditions from the extended relatives (consisting of both FDR+SDR).

## 2. Results

### 2.1. Psychiatric symptoms and syndromes in the participants

Among 1406 individuals, 773 (55%) had one of the five psychiatric diagnoses under the study (Table 1). From among this set, 114 (15%) had two or more of the five syndromes in their lifetime. The remaining 633 (45%) individuals who participated from these families did not have any of these five diagnoses. But, 82 (13%) had “other psychiatric disorders” (Table 1). From among all persons with a DSM-IV diagnosis (773+82=855), 206 (24%) had lifetime co-occurrence of two (or more) DSM-IV syndromes.

**Table 1:** Diagnostic description of participants.

|  | Number of individuals | Percentage |
| --- | --- | --- |
| **Total Participants\*** | **1406** |  |
| **Affected (with any of 5 disorder) Participants** | **773** | **100.00%** |
| SUD | 165 | 21.34% |
| BD | 151 | 19.53% |
| Schizophrenia | 150 | 19.41% |
| OCD | 117 | 15.14% |
| AD | 76 | 9.83% |
| Total Complex SMI | 114 | 14.75% |
| **Complex SMI** |  |  |
| OCD + Schizophrenia | 27 | 23.68% |
| BD + SUD | 22 | 19.3% |
| Schizophrenia + SUD | 20 | 14.91% |
| SAD | 17 | 14.91% |
| BD + OCD | 12 | 10.53% |
| AD + SUD | 6 | 5.26% |
| BD + OCD + SUD | 2 | 1.75% |
| OCD + SAD | 2 | 1.75% |
| OCD + SUD | 2 | 1.75% |
| SAD + SUD | 2 | 1.75% |
| AD + BD | 1 | 0.88% |
| AD + Schizophrenia | 1 | 0.88% |
| **Participating relatives of affected individuals (without any of the 5 disorder)** | **633** | **100.00%** |
| **No Psychiatric disorder (includes Nicotine dependence)** | **551** | **87.05%** |
| **Other psychiatric disorders** | **82** | **12.95%** |

| Other Psychiatric disorders |  |  |
| --- | --- | --- |
| RDD | 16 | 19.51% |
| Depression | 14 | 17.07% |
| Alcohol Abuse | 10 | 12.2% |
| Dysthymia | 10 | 12.2% |
| Generalized Anxiety Disorder | 10 | 12.2% |
| Adjustment Disorder | 6 | 7.32% |
| Agoraphobia | 3 | 3.66% |
| Panic Disorder | 3 | 3.66% |
| ADHD | 2 | 2.44% |
| Generalized Anxiety Disorder + Alcohol Abuse + OCPD | 1 | 1.22% |
| Depression + Alcohol Abuse | 1 | 1.22% |
| Depression + Dysthymia | 1 | 1.22% |
| Dissociation | 1 | 1.22% |
| Dysthymia + Anxiety | 1 | 1.22% |
| Mild Cognitive Impairment | 1 | 1.22% |
| Primary Insomnia | 1 | 1.22% |
| Social Phobia | 1 | 1.22% |

SUD- Substance use disorder; BD- Bipolar Disorder; OCD- Obsessive compulsive disorder; AD- Alzheimer's dementia; SMI- Severe mental illness; SAD- Schizoaffective disorder; ADHD- Attention Deficit Hyperactivity Disorder; RDD – Recurrent Depressive Disorder; Depression – Major Depressive Disorder, Single episode; OCPD – Obsessive Compulsive Personality Disorder.

The sample was densely represented by participants from south-India, with around 12% of participants from other parts of India (Supplementary figure S1). A high socio-functional disruption was noted in terms of remaining single (unmarried/ separated) (38%) and unemployed (20%) with an average illness duration of more than a decade (14±12 years) in the affected participants (Demographic details in Supplementary Table S1).

DSM-5 cross-cutting symptom measure revealed that only a third (34%) of the sample had no psychiatric symptom above the clinical frequency threshold. While 20% had one symptom, four participants had ten out of thirteen clinically significant domain symptoms (for the entire range across each group see figure S2). Forty percent of the unaffected relatives had a minimum one significant psychiatric symptom. Overall, in the study participants, depressive symptoms (26%) were the most common reported symptom followed by somatic symptoms (24%), substance use (23%), anxiety (20%) and sleep (20%). Suicidal ideation was present in 11% of the individuals in the last two weeks. Dissociative symptoms (3%), mania (6%) and psychotic symptoms (8%) were cross-sectionally the least prevalent domains. Other symptoms like memory problems, anger problems, obsessive-compulsive symptoms, and personality dysfunction were detected in more than one-tenth of the individuals (Figure 1). Patients with SUD, OCD, and AD had respective symptom domains (87%, 91% & 87%, respectively) as the most prevalent. Depression was the most prevalent clinical symptom in the prior two-weeks in patients with BD (22%), SCZ (33%), complex SMI (43%) and other psychiatric disorders (41%) groups (Figure S2).

**Figure 1:**
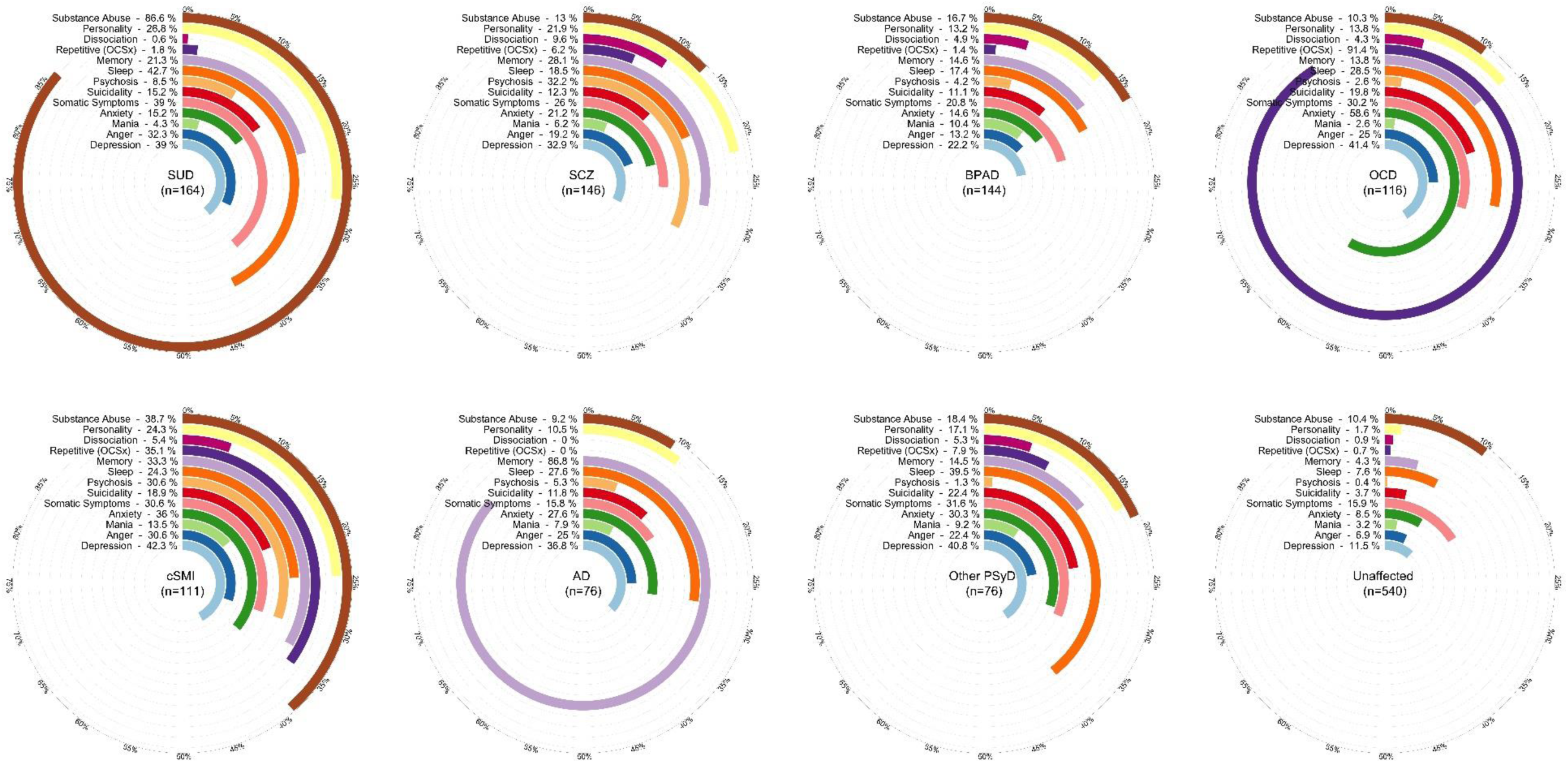
Point Prevalence of psychiatric symptoms in participants across diagnostic categories as measured by DSM5 level-1 cross-cutting measure. various domains of psychiatric symptom crossing the threshold for clinical significance are observed in participants across all diagnostic categories. A significant proportion of the apparently unaffected participants also have different sub-syndromal clinical symptoms. This indicates the phenotypic diversity within the clinical categories. SUD- Substance use disorder; BPAD- Bipolar Affective disorder, OCD- Obsessive-Compulsive disorder; SCZ- Schizophrenia; AD- Alzheimer’s Dementia; cSMI- Complex Severe Mental Illness, Other PsyD- Other Psychiatric Disorders.

### 2.2. The pattern of aggregation of psychiatric diagnosis within the families

The 481 multiplex families in the study have a median of 3 (range: 1-14, mean-2.9, SD-1.3) individuals participating from each family. Families with differing syndromes in affected individuals were more frequent (61%, 295 families). In the other 186 families, all affected individuals had the same diagnosis. SUD families were the majority with 49% (92 families) of all families with similar disorders, and each of the remaining four diagnoses had only 10%-15% share (Figure 2a). The majority of participants with SUD (68%) were from families with similar diagnoses, but this proportion was lower in other diagnoses (AD-38%, OCD-39%, SCZ-27%, BD-25%) (Figure 2b).

**Figure 2:**
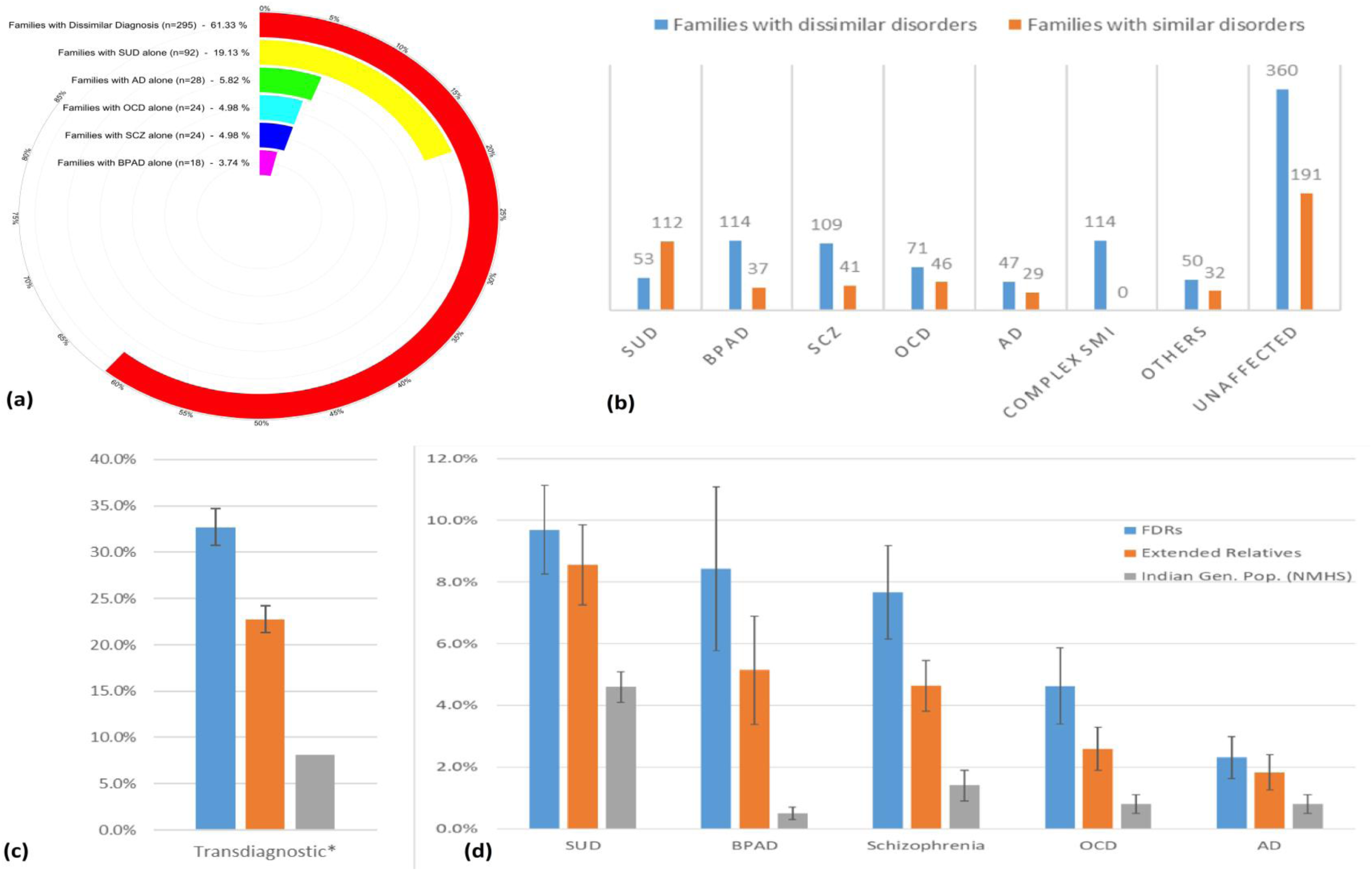
Frequency of occurrence of psychiatric syndromes in multiplex families. (a) The frequency of families with multiple affected members with similar and dissimilar diagnoses are depicted. Most of the families had more than one diagnostic phenotype (dissimilar diagnoses). Half of all the families with one type of diagnosis consisted of only SUD. (b) Participants in all different diagnostic categories had different proportions of families with more than one diagnostic phenotype (dissimilar diagnoses). Participants of all diagnostic categories, except for SUD, hailed from families with multiple diagnostic phenotypes. (c) Prevalence of any the five disorders is around 4 times higher in the first-degree relatives and 3 times higher in extended relatives (combined first- and second-degree relatives) of the study participants compared to the general population prevalence. *Transdiagnostic prevalence in the general population is calculated by the sum of the prevalence rates of all five disorders in the general population. (d) Prevalence of BPAD in FDRs of the participant from the multiplex family was nearly 17 times, schizophrenia and OCD-6 times, AD-3 times and SUD-2 times higher than that of the general population. Similarly, extended relatives had 2-10 times higher prevalence (2% with AD, 3% with OCD, 5% with BPAD and SCZ, and 8% with SUD) of these disorders in comparison to lifetime prevalence in general population. Transdiagnostic-Overall prevalence of any of the five disorders, SUD- Substance use disorder; BPAD- Bipolar Affective disorder, OCD- Obsessive-Compulsive disorder; AD- Alzheimer’s Dementia; Complex SMI- Complex Severe Mental Illness; Others-Other psychiatric disorders, Indian Gen.Pop. - Indian general Population prevalence (based on National mental health survey, Gururaj et al., 2016 and Mathuranath et al., 2010).

The lifetime prevalence of psychiatric syndromes was calculated among relatives (SDRs=19,559, 14.18±5.45 per participant, FDRs=9,241, 6.7±3.18 per participant) of 1379 participants from 477 families. Though one another affected FDR was the minimum inclusion criteria, an average of 2.12 (±1.35) FDRs and an additional 1.05 (±1.66) SDR of the affected participants had one of the five conditions. Twenty-seven individuals were excluded due to insufficient FIGS data. The overlapping relatives within the families are adjusted for double-counting using complex sample statistics, as described in the methods section. On estimating the prevalence, 33% (95% CI: 31%-35%) of FDRs and 23% (21%-24%) of extended relatives of the participants had any of the five diagnoses. This implies that every one in three FDR and one in the fifth SDR of a person from multiplex families was affected with any of the five disorders. A similar trend was observed in FDRs for all five disorders [SUD: 10% (CI:8%-11%), BD: 8% (6%-11%), SCZ: 8% (6%-9%), OCD: 5% (3%-6%) and AD: 2% (2%-3%)]. The same order of frequency was noted even after adding SDRs though prevalence was lesser than that in FDRs alone (Figure 2C,2D & Table 2).

**Table 2:** Prevalence rates of similar and dissimilar psychiatric disorders and prevalence ratios across diagnostic categories.

|  | FDRs |  |  |  |  | FDR+SDRs |  |  |  |  |
| --- | --- | --- | --- | --- | --- | --- | --- | --- | --- | --- |
|  | P<sub>Similar</sub><br>diagnosis | P<sub>Dissimilar</sub><br>diagnosis | P<sub>Similar</sub> diagnosis<br>among affected | P<sub>Dissimilar</sub><br>diagnosis among<br>affected | PR | P<sub>Similar</sub><br>diagnosis | P<sub>Dissimilar</sub><br>diagnosis | P<sub>Similar</sub><br>diagnosis among<br>affected | P<sub>Dissimilar</sub> diagnosis<br>among affected | PR |
| **SUD** | 25.91% | 6.42% | 0.80 | 0.20 | 4.04 | 22.89% | 3.85% | 0.86 | 0.14 | 5.94 |
| **BD** | 24.72% | 12.03% | 0.67 | 0.33 | 2.05 | 13.01% | 8.63% | 0.60 | 0.40 | 1.51 |
| **Schizophrenia** | 20.13% | 11.83% | 0.63 | 0.37 | 1.70 | 9.35% | 7.33% | 0.56 | 0.44 | 1.28 |
| **OCD** | 20.73% | 10.71% | 0.66 | 0.34 | 1.94 | 10.24% | 8.76% | 0.54 | 0.46 | 1.17 |
| **AD** | 11.41% | 8.24% | 0.58 | 0.42 | 1.38 | 10.60% | 7.31% | 0.59 | 0.41 | 1.45 |

SUD- Substance use disorder; BPAD- Bipolar Affective disorder, OCD- Obsessive Compulsive disorder; AD- Alzheimer's Dementia; Complex SMI- Complex Severe Mental Illness.

The prevalence of affected FDRs was least for participants with AD at 20% (16%-23%) and highest for BD at 37% (28%-45%) (supplementary table S2). Affected participants had a higher prevalence of the same disorder in their relatives (11%-26% in FDRs, 9%-23% in extended relatives) than the other four disorders put together (6%-12% in FDRs, 4%-9% in SDRs). This suggests that similar diagnoses do aggregate in the multiplex families. However, the occurrence of other disorders was also quite significant. Nearly every third affected FDR of the participants with AD, SCZ, OCD, and BD had a different diagnosis (Table 2).

## 3. Discussion

Kessler et al.^43^ observed that “*Although mental disorders are widespread, serious cases are concentrated among a relatively small proportion of cases with high comorbidity*”. The “serious cases” in this study were selected from a large tertiary psychiatric hospital after screening for the presence of a strong family history to achieve the project objectives^27^. The five disorders (SUD, BD, SCZ, OCD, and AD) opted in the project cause significant disability and impose a burden on the individuals, their families and the society. A high rate of unemployment and remaining single/separated highlights the social decline as a consequence of psychiatric disorders in these families.

The identified participants had a high familial risk, with nearly 1/3^rd^ of FDRs affected. The majority of multiplex families had members with dissimilar diagnoses and not just the same diagnosis. A good proportion of affected participants had co-occurring syndromes over their lifetime. Diverse cross-cutting symptoms, not restricted to the index syndrome, are also observed in participants. Symptoms were observed in a significant proportion of apparently unaffected FDRs.

### 3.1. Phenotypic diversity: symptom overlap across diagnostic groups

A vast amount of phenotypic diversity is observed within and across individuals. As much as 15% of the individuals with these five psychiatric disorders had co-occurrence of two or more of these syndromes, during their course of illness. If we include any DSM-IV diagnosis, the number goes up to 24%. Even in those without a comorbid diagnosis, the subsyndromal psychiatric symptoms were found across nosological categories^15^. Overall one in every four participants having significant depressive symptoms and 10% having suicidality in the preceding two weeks echoes the severity of psychological problems in such families. Although most of the patients with varied diagnoses were on treatment and in different stages of recovery, the cross-sectional evaluation of psychiatric symptoms in the prior two weeks suggests the presence of a gamut of behavioral and psychological phenotypes. Symptoms transcending across traditional diagnostic boundaries highlight the need for a symptom-based approach ^44-48^ and a dimensional approach suggested by the RDoC^19, 49^.

### 3.2. Clinical status of “unaffected” FDRs

A significant proportion (12.4%) of FDRs who were included in the study for their at-risk status were found to have a diagnosable psychiatric diagnosis on structured clinical evaluation. Among the remaining relatives without any psychiatric morbidity, 39% had a psychiatric symptom crossing the threshold of clinical significance in the DSM-V cross-cutting symptoms measure. A higher proportion of symptoms and psychiatric syndromes in the apparently unaffected FDRs could indicate their higher biological vulnerability^35^ or high psychosocial burden in caring for multiple relatives with SMI^50-52^. This further necessitates long-term follow up of such individuals to understand the biological and psychosocial interactions associated with the evolution of severe phenotypes, as envisaged in ADBS.

### 3.3. Familial aggregation pattern of psychiatric diagnoses

The ascertainment of higher familial risk in this sample is intended to identify ‘genetically enriched’ pedigrees. High familial risk in this sample was demonstrated by the presence of psychiatric disorders in nearly one-third of FDRs and nearly 22% of extended relatives of the participants. A recent national mental health survey (NMHS) in India indicates a lifetime prevalence of SUD (4.6%), BD (0.5%), OCD (0.8%), and SCZ (1.4%) in adult general population^9^. Maximum reported Indian prevalence of AD is 3.77% in those aged >55 years which equates to around 0.8% of the Indian adult population^53^. Thus, the combined prevalence rates of these disorders in the relatives of the participants were 3-4 times greater than that in the general population.

Increased familial risk is the major contributor to the propensity to develop severe mental illnesses^54, 55^. The majority of the families of the participants had different diagnoses among their members than a single specific syndrome (295 vs 196 families). Such diverse diagnoses within families are reported earlier^15, 34, 56-59^. Thus, these transdiagnostic multiplex families may represent a real-world scenario much better than syndrome specific multiplex families. However, similar diagnoses did tend to aggregate within the relatives. The prevalence of similar diagnoses was 1.5 to 4 times that of a dissimilar diagnosis, the highest s being seen in SUD. The higher relative prevalence of similar diagnosis acknowledges, though indistinct, the boundaries in existing diagnostic categorization. This indicates a need to design etiological studies within this transdiagnostic framework^4, 59-62^.

### 3.4. Implications

Transdiagnostic research is yielding novel insights into role of the neuronal markers^8, 10, 12, 20, 24, 63^, molecular^7, 64, 65^, genetic^3, 5, 7, 22, 34, 54, 55^ as well as psychosocial factors^13, 66^. Pleiotropic genetic loci associated with multiple psychiatric conditions are being identified^67^. An understanding of mechanisms that are unique as well as shared across disorders considering interindividual and intrafamilial phenotypic diversity is thus necessary^18, 19, 49^. Consenting individuals from this transdiagnostic multiplex families undergo detailed assessment for developmental anomalies, temperament, personality, past psychosocial adversities, socioeconomic status, and the severity of psychopathology. Structural and functional magnetic resonance imaging (MRI), electroencephalography/evoked potential(EEG), functional near infra-red spectroscopy(fNIRS) and eye movement tracking are being done for these participants since August 2017^27^. A repository of induced pluripotent stem cells, lymphoblastoid cell lines and other biomaterials (serum, plasma, DNA) has been established and is being expanded. The bio-repository comprised of this “enriched” sample will be maximally representative of individuals with severe psychiatric disorders and those with very high susceptibility; and, thus, provide a great opportunity to answer several questions related to the neurobiology and pathogenesis of psychiatric disorders. This cohort is planned to undergo biennial assessments with detailed clinical, neuroimaging and neurophysiological tools, the second phase of which has begun in August 2019.

ADBS intends to make the raw de-identified data (clinical, MRI, EEG, fNIRS, eye tracking and DNA sequence data) and biological resources (plasma samples, genomic DNA and cell lines) generated from its research activities available to the wider scientific community. A data bank and bio-repository are being developed in the project. All raw data from clinical endophenotyping studies will be available for sharing with external investigators 12 months after the baseline data acquisition is completed. All cell lines generated by ADBS will be made available for sharing with external investigators, without the need to collaborate with an ADBS investigator, 12 months after the generation of cell lines has been completed from a single-family. Even before this embargo period, external investigators can embark on a formal collaboration with ADBS investigators to use the data and resources. The sharing of data and resources will follow due process with approval from the resource sharing and management boards. A full list of ADBS PIs and their contact details are available on the ADBS websites (https://ncbs.res.in/adbs/home & http://adbsnimhans.org/).

### 3.5. Limitations

Few limitations are to be acknowledged. The transdiagnostic approach is limited to the affected status of five specific psychiatric disorders. This a major step in using a transdiagnostic approach in understanding the biological bases of severe mental disorders; but is inadequate to interrogate mechanisms of common syndromes of anxiety and depressive disorders^14^. The five syndromes become manifest from young adult life to late life (AD). A uniform age-at-risk of 13 years and above may not be equally representative of each of the syndromes. A non-probabilistic sampling method was applied for the efficient identification of highly vulnerable families. Hence, we need to acknowledge the possibility of selection bias and limitations in generalizing the estimates to the wider universe of multiplex families. The affected status of non-participating family members is ascertained by family history methods, and thus the knowledge of mildly affected members or those who chose not to disclose their family history may have been missed. However, family history was obtained from at least two FDRs that would partially compensate for any possible reporting bias. An unbiased ascertainment for non-occurrence is not possible in relatives given the persistent risk for future occurrence. Nevertheless, the obtained estimates are conservative and the actual familial risk might be much higher than reported.

### 3.6. Conclusions

The ADBS study is congregating participants from extremely dense families with five psychiatric disorders. Most of these families have their members unrestricted to a single-diagnosis with a high prevalence of co-occurrence of syndromes in the affected individuals. In addition, a significant proportion of participants, across all diagnostic groups (including the unaffected relatives) have subsyndromal psychiatric symptoms of diverse domains. However, similar syndromes aggregate within the families and specific symptoms aggregate within individuals with particular disorders. This sample would thus be helpful in elucidating the pathogenetic mechanisms that are specific and shared across disorders.

## Data Availability

Data will be available on reasonable request.

## Funding acknowledgement

This research is funded by the Accelerator program for discovery in brain disorders using stem cells (ADBS) (funded by the Department of Biotechnology, Government of India (BT/PR17316/MED/31/326/2015)).

---

**Table S1:**
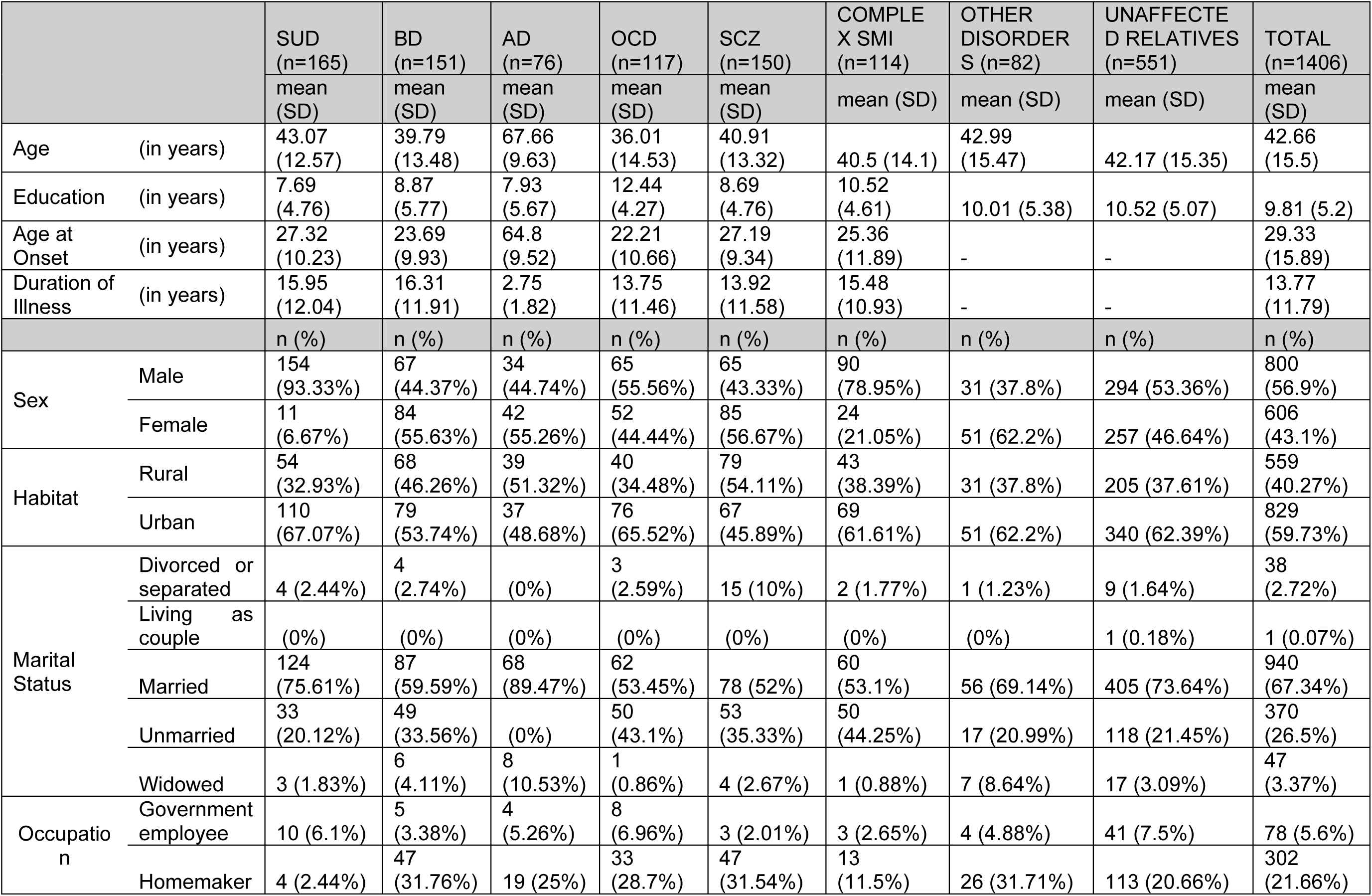
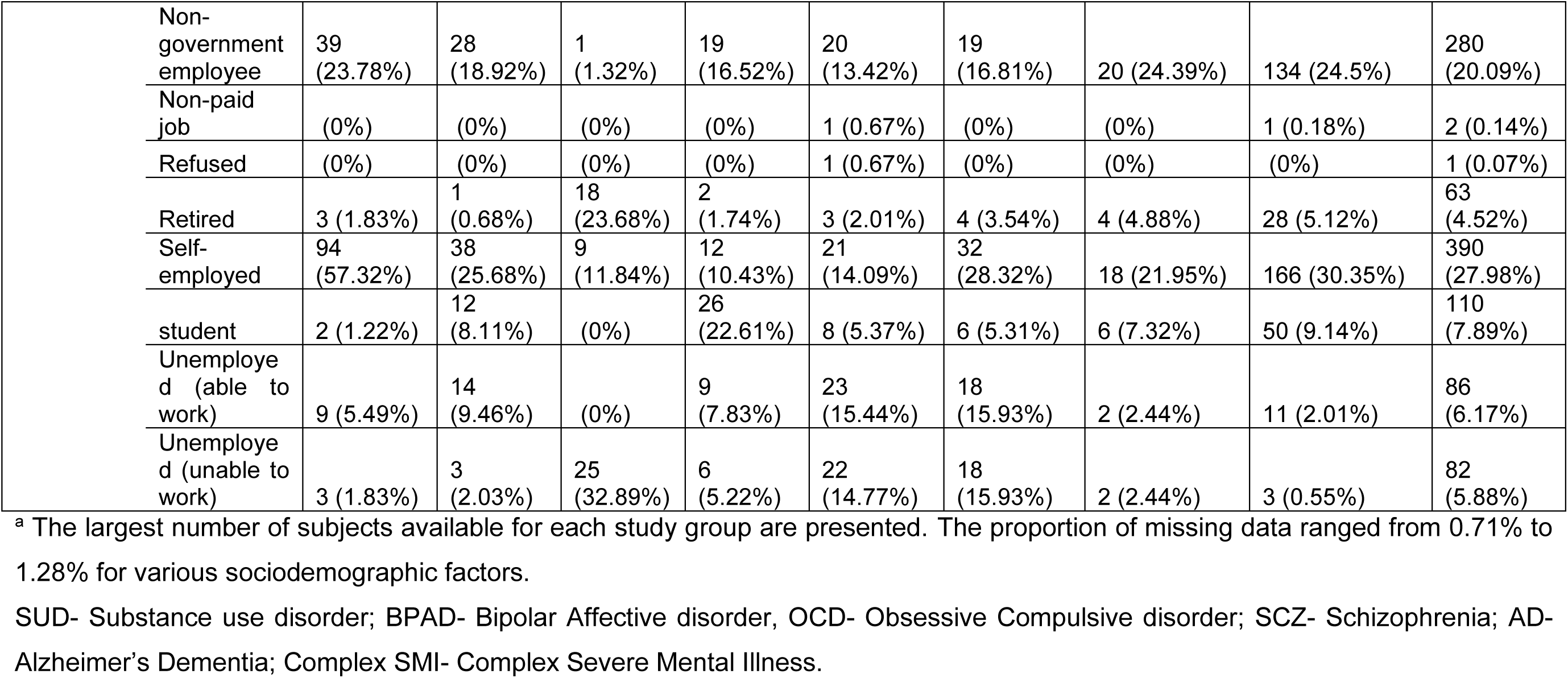
Baseline characteristics (continuous variables) of participants across different diagnostic categories^a^.

**Table S2:** Prevalence rates of the five psychiatric disorders in the FDRs as well as FDRs and SDRs of individuals from multiplex families across diagnostic categories.

| Diagnosis in Participants | **P<sub>SUD</sub>** |  | **P<sub>BD</sub>** |  | **P<sub>AD</sub>** |  | **P<sub>OCD</sub>** |  | **P<sub>Schizophrenia</sub>** |  | **P<sub>Transdiagnosis</sub>** |  |
| --- | --- | --- | --- | --- | --- | --- | --- | --- | --- | --- | --- | --- |
|  | FDR | Extended Relative S | FDR | Extended Relative S | FDR | Extended Relative S | FDR | Extended Relative S | FDR | Extended Relative S | FDR | Extended Relative S |
| **SUD** | 25.91% | 22.89% | 0.56% | 0.56% | 3.15% | 1.88% | 1.56% | 0.85% | 0.65% | 0.56% | 32.33% | 26.75% |
| **BD** | 5.00% | 4.20% | 13.01% | 13.01% | 5.54% | 3.42% | 1.49% | 0.92% | 0.00% | 0.10% | 36.75% | 21.64% |
| **Schizophrenia** | 6.27% | 4.62% | 1.90% | 1.92% | 20.13% | 9.35% | 1.68% | 0.73% | 0.19% | 0.08% | 31.95% | 16.68% |
| **OCD** | 4.15% | 4.54% | 1.80% | 1.63% | 3.80% | 2.17% | 20.73% | 10.24% | 0.30% | 0.26% | 31.44% | 18.99% |
| **AD** | 4.80% | 4.22% | 1.00% | 1.00% | 1.68% | 1.57% | 0.61% | 0.52% | 11.41% | 10.60% | 19.66% | 17.90% |
| **Complex SMI** | 13.68% | 10.49% | 5.36% | 5.89% | 4.75% | 3.29% | 6.76% | 3.41% | 1.77% | 1.51% | 33.42% | 24.06% |
| **Other Psychiatric Disorder** | 10.29% | 8.21% | 5.95% | 5.21% | 9.13% | 4.86% | 4.78% | 2.34% | 1.00% | 0.58% | 35.77% | 21.94% |
| **Unaffected** | 9.14% | 8.78% | 5.26% | 5.21% | 8.21% | 5.35% | 4.89% | 2.69% | 3.10% | 3.09% | 33.35% | 25.17% |
| **Total** | 9.69% | 8.57% | 5.14% | 5.09% | 7.66% | 4.64% | 4.63% | 2.59% | 2.31% | 1.83% | 32.72% | 22.76% |

**Figure S1:**
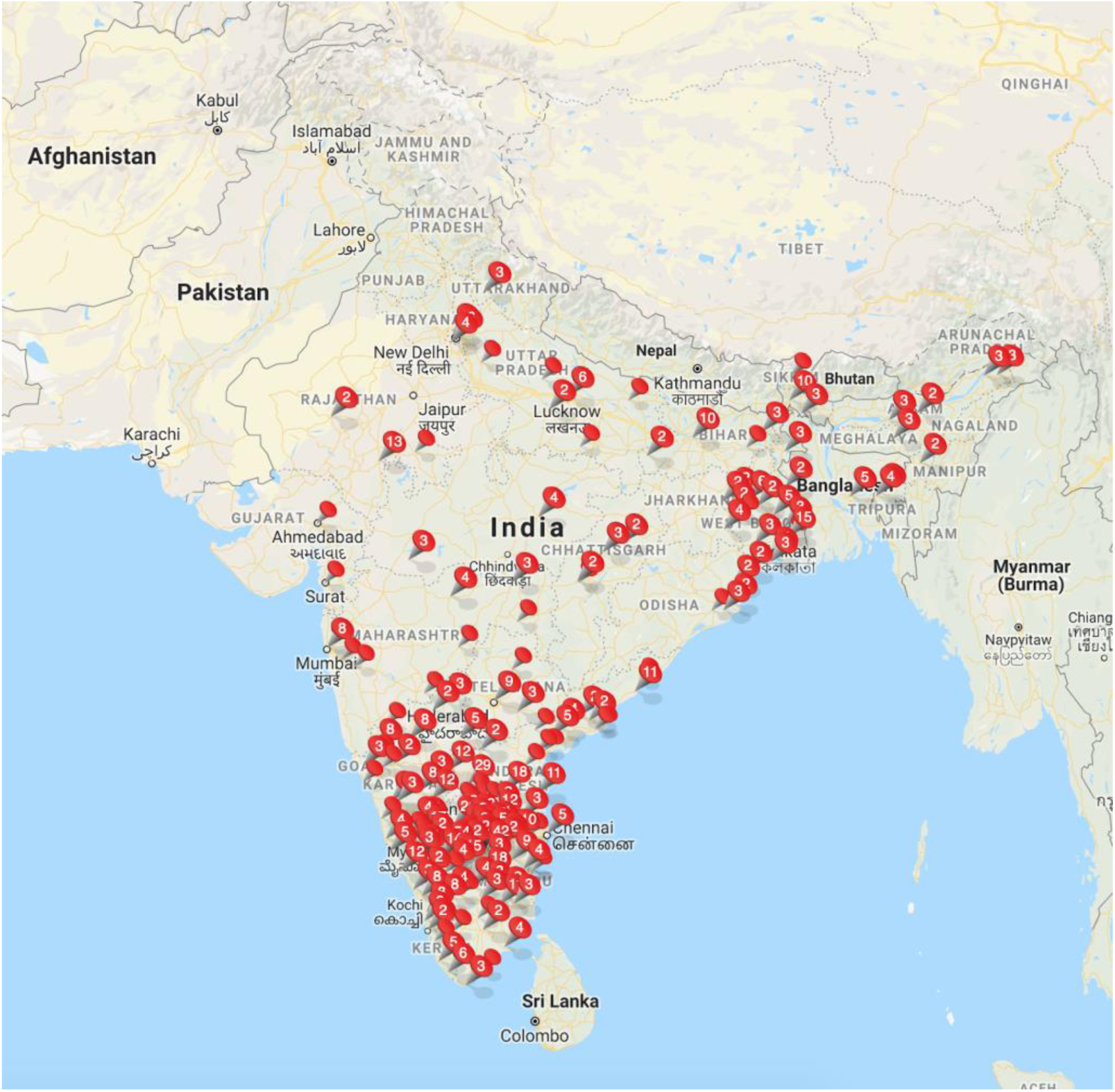
Geographical distribution of the study participants. The sample has a higher representation of the population from 5 southern states of India, primarily due to the proximity of the study site.

**Figure S2:**
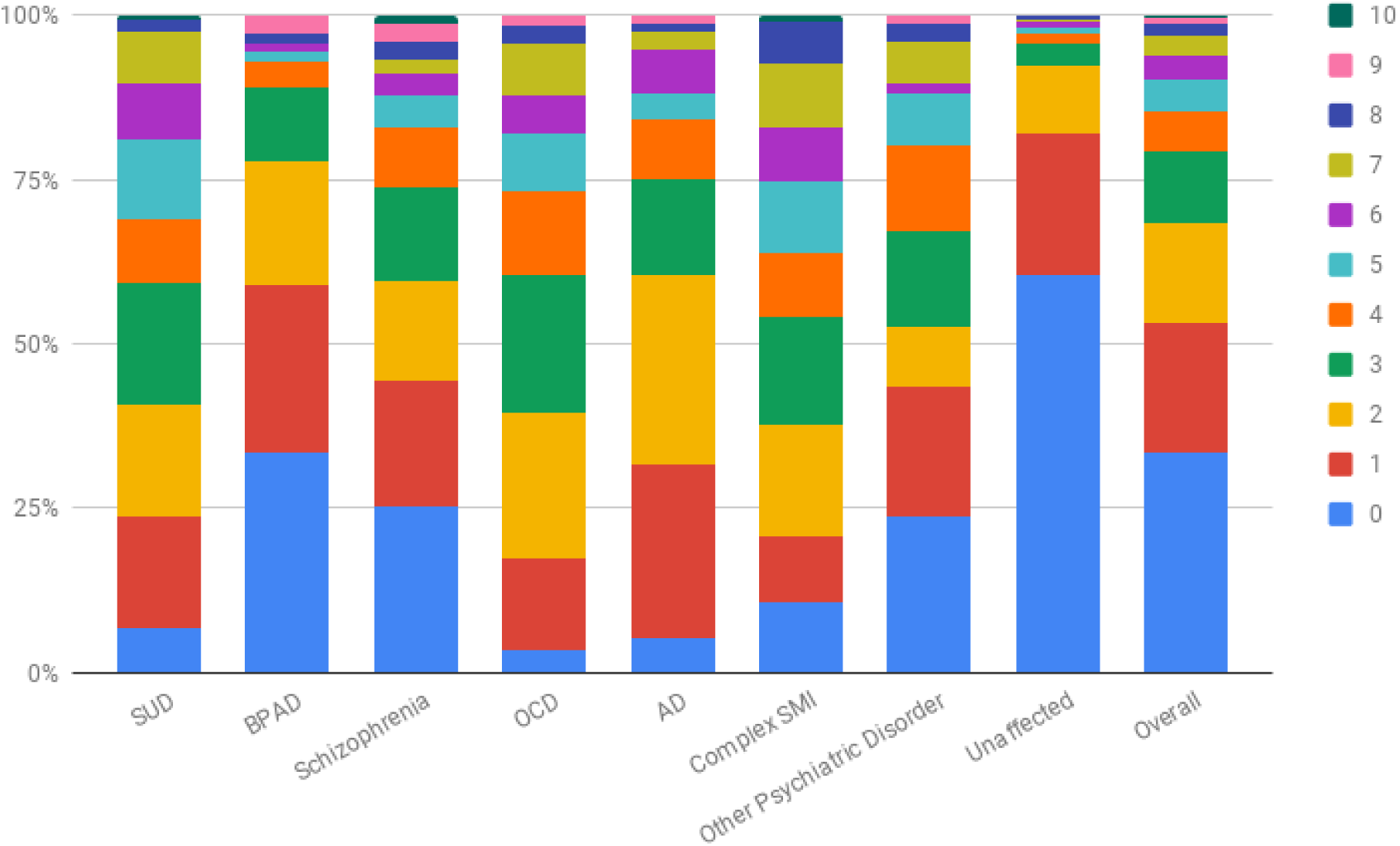
100% stacked column of distribution of psychiatric symptoms counts in participants across diagnostic categories as measured by DSM5 level-1 cross-cutting measure. Each stack represents the number of symptom domains screened positive in DSM5 cross-cutting symptom measure. Less than 30% of the study participants have no clinically significant symptom. But nearly 50% have 2 or more domains of symptoms in the last 2 weeks displaying the intra-individual phenotypic diversity. SUD- Substance use disorder; BPAD- Bipolar Affective disorder, OCD- Obsessive Compulsive disorder; AD- Alzheimer’s Dementia; Complex SMI- Complex Severe Mental Illness.

